# Gene Therapy Restores the Transcriptional Program of Hematopoietic Stem Cells in Fanconi Anemia

**DOI:** 10.1101/2021.07.20.21260460

**Authors:** Miren Lasaga, Paula Río, Amaia Vilas-Zornoza, Nuria Planell, Susana Navarro, Diego Alignani, Beatriz Fernández-Varas, Josune Zubicaray, Roser M. Pujol, Eileen Nicoletti, Jonathan D. Schwartz, Julián Sevilla, Marina Ainciburi, Asier Ullate-Agote, Jordi Surrallés, Rosario Perona, Leandro Sastre, Felipe Prosper, David Gomez-Cabrero, Juan A. Bueren

## Abstract

Fanconi anemia (FA) is an inherited disease associated with marked hematopoietic stem and progenitor cell (HSPC) defects. Ongoing clinical trials have shown that lentiviral-mediated gene therapy can ameliorate bone marrow failure (BMF) in non-conditioned FA patients thanks to the proliferative advantage of corrected FA HSPCs. Here we investigated whether gene therapy can revert affected molecular pathways in diseased HSPCs, a question not previously addressed in any HSC gene therapy trial. Single-cell RNA sequencing was performed in chimeric populations of corrected and uncorrected HSPCs coexisting in BM of gene therapy treated FA patients. Our study demonstrates that gene therapy reverts the transcriptional signature of FA HSPCs, which then resembles the transcriptional program of healthy donor HSPCs. This includes a downregulated expression of TGF-β and p21, typically upregulated in FA HSPCs, and upregulation of DNA damage response and telomerase maintenance pathways. Our results show for the first time the potential of gene therapy to rescue defects in the HSPC transcriptional program from patients with inherited diseases, in this case in FA characterized by BMF and cancer predisposition.

## INTRODUCTION

The potential of gene therapy (GT) to correct a variety of human inherited diseases, including primary immunodeficiencies and hemoglobinopathies has been well demonstrated in previous clinical studies (Ferrari, Thrasher et al., 2021). Additionally, we have recently shown that lentiviral-mediated GT can also enable correction of more complex diseases, such as Fanconi anemia (FA), in which marked phenotypic defects are already evident in the self-renewing HSCs. As previously reported, the infusion of corrected CD34^+^ cells in non-conditioned FA patients resulted in a marked HSPC proliferative advantage, which facilitates the progressive increase in the number of bone marrow (BM) and peripheral blood (PB) gene-corrected cells (Rio et al., 2019).

Despite significant advances in the field of GT, the full potential of this therapeutic modality will depend on its capacity to reestablish the molecular circuits and the functional potential of diseased cells. This is even more relevant in syndromes such as FA, characterized by DNA repair defects resulting in progressive accumulation of DNA damage, and additional molecular responses triggered by a defective FA/BRCA pathway. In patients treated in the FANCOLEN-I clinical trial, the presence of a chimeric population of corrected and uncorrected FA HSPCs, none of them exposed to any conditioning agent, let us comparatively investigate the differential molecular pathways that characterize each of these populations coexisting in the BM of these patients (**Fig.1**). This approach allowed us to demonstrate that lentiviral-mediated GT not only mediates a progressive engraftment of FA patients with gene corrected HSPCs, but also results in the reprogramming of the transcriptional signature of FA HSPCs, which then resembles the molecular circuits of healthy HSPCs, thus accounting for the phenotypic correction of FA HSPCs.

## RESULTS

### Gene therapy modifies the HSPC transcriptional program in FA patients

Bone marrow (BM) CD34^+^ cells from four FA-A patients (FA-02002, FA-02004, FA-02006, and FA-02008) who had respectively been treated with gene therapy 5, 4, 3, and 2 years previously, were purified and analyzed by single-cell RNA sequencing (scRNAseq), as described in **Fig 1**. qPCR analyses from these samples showed the presence of 0.77 vector copy numbers (VCN) per cell in the BM of patient 02002, and 0.45, 0.26, and 0.29 VCNs per cell in purified CD34^+^ cells in patients FA-02004, FA-02006, and FA-02008 respectively (see Materials and methods). Since the average VCN per corrected cell was of 1.0 copies(Rio, Navarro et al., 2019), the proportion of corrected cells in these patients is estimated to be 77%, 45%, 26%, and 29%, respectively, revealing the chimeric nature of the HSPC populations residing in their BM.

**Figure 1.**
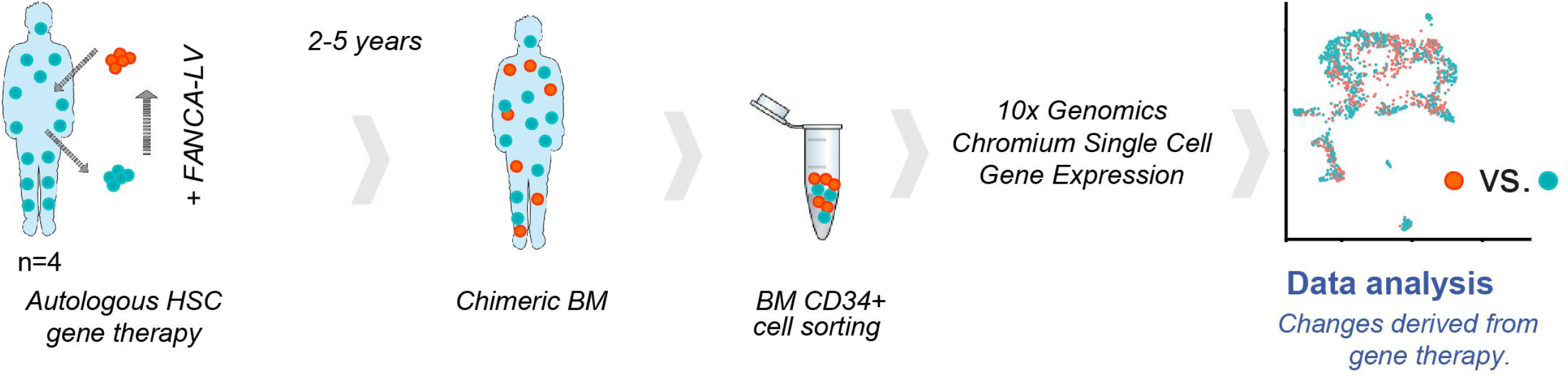
Experimental design of scRNAseq analyses performed in FA-A patients 2-5 years after lentiviral-mediated gene therapy. Four FA-A patients who had been treated by *ex vivo* lentiviral-mediated gene therapy in the absence of conditioning were included in this study. At 2-5 years post-gene therapy, these patients harbored a chimeric population of corrected and uncorrected hematopoietic stem and progenitor cells (HSPCs) in their bone marrow (BM). Aliquots of BM CD34^+^ cells from these patients were sorted and processed for single-cell RNA-seq. Bioinformatic analyses were then conducted to comparatively investigate changes in the transcriptional program of corrected *versus* uncorrected HSPCs, coexisting in each of the gene therapy treated patients.

CD34^+^ cells from FA patients were classified according to transcriptional profiles previously described(Velten, Haas et al., 2017, Zheng, Terry et al., 2017), which identified twelve different HSPC clusters that corresponded to primitive HSCs and more committed lympho-hematopoietic progenitor cells (See analyses from patient FA-02006 in **Fig. 2A;** and patients FA-02002, FA-02004 and FA-02008 in **Fig. S1A, B, C, D)**. To investigate the impact that GT had in the transcriptional program of FA HSPCs, CD34^+^ cell subpopulations from GT-treated FA patients were classified as FANCA^+^ and FANCA^-^, based on the expression of *FANCA* by scRNAseq (see materials and methods). As shown in **Fig. 2B** and **2C**, the presence of FANCA^+^ cells was evident in all HSPC populations represented with at least 30 cells per cluster. No significant differences in the proportion of FANCA^+^ cells present in these subpopulations were observed among the four patients (**Table S1**). Additionally, a wide range of FANCA expression was observed in samples analyzed, although higher expression levels were observed in most HSPC subpopulations from patient FA02002, compared with the other patients **(Fig. 2D)**.

**Figure 2.**
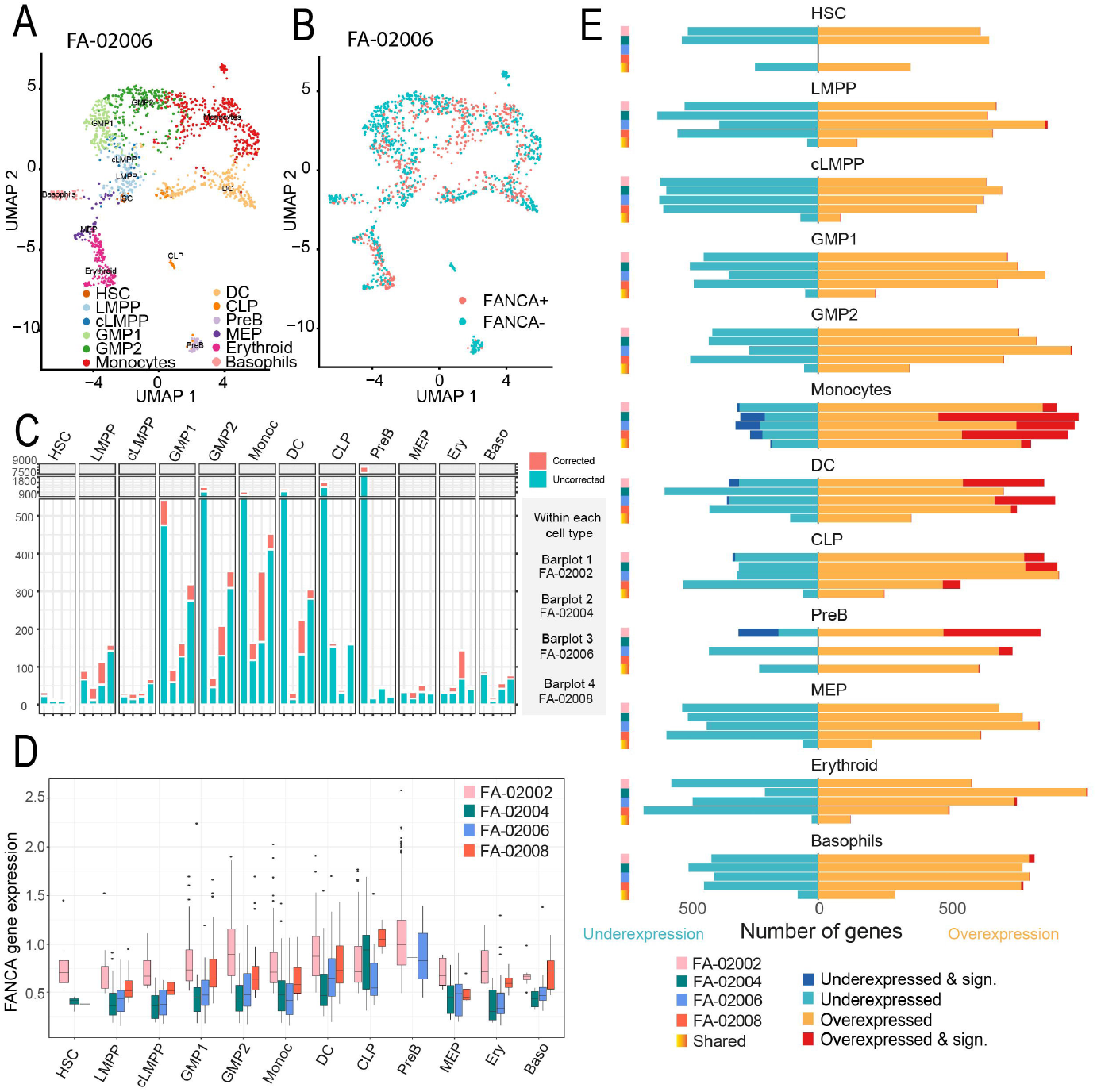
scRNAseq analysis of corrected and uncorrected hematopoietic stem and progenitor cells from FA-A patients 2-5 years after lentiviral-mediated gene therapy. **(A)** UMAP plot showing the clustering analysis for BM CD34^+^ cells from a FA-A patient previously treated by gene therapy (FA-02006 patient as an example; see **Fig. S1** for patients FA-02002, FA-02004 and FA-02008). A total of 12 clusters were identified, spanning the different HSPC subpopulations. Identified clusters include an HSC cluster (hematopoietic stem cell; brown). Clusters with megakaryocytic-erythroid identity include MEP (erythroid-megakaryocyte progenitor; purple), Erythroid (erythroid progenitor; pink), and Basophils (basophil progenitor; light pink). Clusters with lympho-myeloid identity include LMPP (lymphoid-primed multipotent progenitor; light blue), Cycling-LMPP (blue), CLP (common lymphoid progenitor; orange), GMP1 and GMP2 (granulocyte-monocyte progenitor; light green and green), Monocytes (monocyte progenitor; red), DC (dendritic cell progenitor; nude), and PreB (B cells progenitor; light purple). **(B)** The same UMAP as shown in panel A, highlighting the distribution of FANCA^+^ cells (FANCA mRNA detectable; red) versus FANCA^-^ cells (FANCA mRNA levels are below detection limit; blue). **(C)** Barplot showing the total number of cells in the different HSPC populations corresponding to the four gene therapy treated patients. In each case, the number of FANCA^+^ (red) and FANCA^-^ (blue) cells is shown and the percentage represented by FANCA^+^ is written in the top of the bar. **(D)** Boxplot of integrated and normalized FANCA gene expression in FANCA^+^, depicted by cell type and FA individual (n=3). **(E)** Barplot representation of the differential expression analysis between FANCA^+^ and FANCA^-^ for each FA patient (n=4) and for each of the HSPC populations. In each case, the number of upregulated and downregulated genes is shown. Upregulated genes were defined as logFC>0.25 (orange), upregulated and significant genes were defined as a logFC>0.25 and adjusted p-value<0.05 (red), downregulated genes were defined as a logFC<-0.25 (light blue) and downregulated and significant genes were define as logFC<-0.25 and adjusted p-value<0.05 (dark blue). The number of genes with the same behavior (upregulated or downregulated) in the four individuals is shown below each HSPCs (labeled as “*shared*”).

To investigate the impact that ectopic FANCA expression had in the transcriptional program of HSPCs, we performed a differential expression analysis between FANCA^+^ and FANCA^-^ HSPCs present in each GT-treated patient. As shown in **Fig. 2E**, the ectopic expression of *FANCA* upregulated a high number of genes compared to the number of genes that were downregulated. As expected, genes with statistically differential expression corresponded to subpopulations with a higher representation (i.e., monocytic CD34^+^ cells; genes with significantly up-regulated and down-regulated expression (p-value< 0.001) are shown in dark red and dark blue bars, respectively in **Fig. 2E**). Remarkably, most of the upregulated and downregulated genes in FANCA^+^ vs. FANCA^-^ cells showed the same expression pattern in each of the analyzed patients (see fifth bar at the bottom of each patient’s bars in **Fig. 2E**). To select the genes with a robust differential expression between FANCA^+^ versus FANCA^-^ HSPCs, we considered those genes that showed a significant differential expression (abs(logFC)>0.25 and adjusted p-value<0.05) in at least one cell type and in at least three patients. In addition, changes in gene expression should be in the same direction in all the four patients. Based on these criteria, a total number of 152 differentially expressed genes were identified (See **Table S2**).

Taken together, data obtained in these analyses indicate that the ectopic expression of *FANCA* induced long-term reproducible changes in the gene expression program of HSPCs from FA-A patients.

### Gene therapy reprograms the transcriptional signature of FA HSPCs towards the transcriptional profile characteristic of healthy HSPCs

Next, we investigated whether changes in the transcriptional program of gene-corrected FA-A HSPCs resembled profiles characteristic of healthy HSPCs. To this end, scRNAseq data from GT-treated FA CD34^+^ cells was compared with that obtained from HD CD34^+^ cells. As shown in **Fig. 3A** (see also **Expanded View Figure S2**) the ectopic expression of *FANCA* (FANCA^+^ cells) was detected in the different CD34^+^ cell clusters. Additionally, **Fig. 3B** shows that *FANCA* expression levels in the different HSPC subpopulations are highly heterogeneous, not only in the case of GT-treated FA patients, but also in HDs. Despite this heterogeneity, in eight out of the twelve HSPC subpopulations, physiological levels of FANCA mRNA were significantly higher in HD HSPCs compared with the corresponding values observed in FANCA^+^ HSPCs from GT-treated FA patients. This observation is consistent with the average insertion of 1 proviral *FANCA* copy per corrected cell after GT, in comparison with the 2 copies of WT *FANCA* per HD cell, and also with the moderate activity of the phosphoglycerate kinase (PGK) promoter that was selected for the therapeutic vector because of its favorable safety profile.

**Figure 3.**
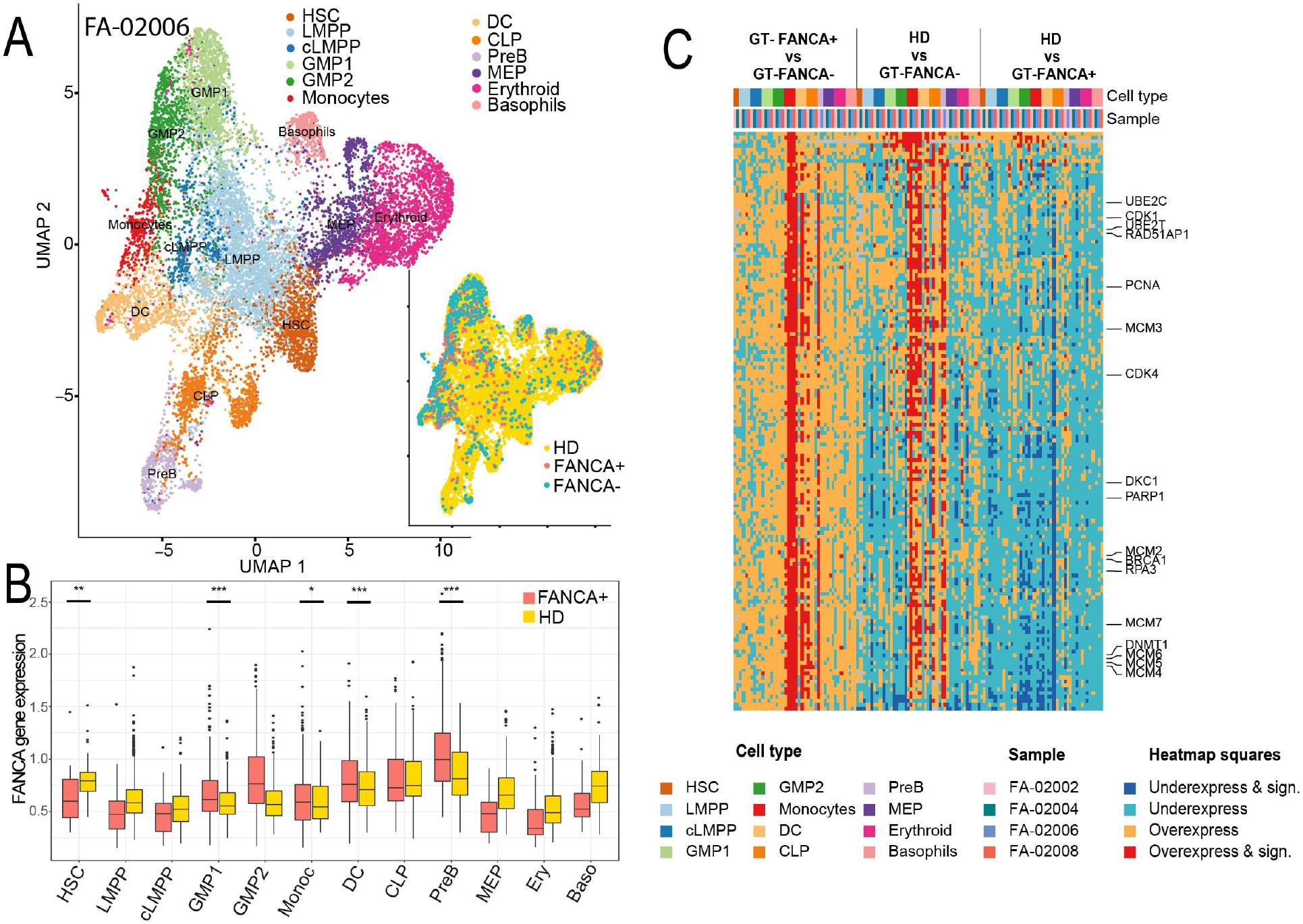
Comparisons of the gene expression signature between corrected and uncorrected HSPCs coexisting in gene therapy treated FA patients. **(A) Left larger panel**: UMAP plot showing the clustering analysis of CD34^+^ BM cells after integration of data from a gene therapy treated FA-A patient and a healthy donor (FA-02006 is included as a representative example; see Fig. S2 for the rest of individuals). **Right smaller panel**: the same UMAP as shown in left panel but highlighting the distribution of HD HSPCs (yellow), FANCA^+^ HSPCs (red) and FANCA^-^ HSPCs (blue). Clusters identification as in Figure 1. **(B)** Boxplot representation of normalized single-cell FANCA expression of FANCA^+^ cells (red) and HD cells (yellow) by HSPC cluster. For each cell type, differences in the expression levels between the ectopic expression of FANCA from corrected FA CD34^+^ cells and expression levels corresponding to HD CD34^+^ cells are shown (*adjusted p-value <0.05; **adjusted p-value < 0.01; ***adjusted p-value < 0.001) **(C)** The figure shows the results associated to three differential expression contrasts: FANCA^+^ vs FANCA^-^ HSPCs from GT-treated FA patients; HD HSPCs vs FANCA^-^ HSPCs from GT-treated patients; and HD HSPCs vs FANCA^+^ HSPCs from GT-treated patients. The second row shows the twelve different CD34^+^ cell types, and the third one the sample identification from each of the four GT-treated patients. Up-regulated genes (logFC>0) are shown in orange and those with significant upregulation in red (logFC>0.25 and adjusted p-value<0.05). Down-regulated genes (logFC<0) are shown in light blue and those with significant downregulation in dark blue (logFC<-0.25 and adjusted p-value<0.05). Unsupervised hierarchical clustering using Pearson distance and average linkage method was applied for gene classification. The genes included in the heatmap are those that for at least one cell type are identified as differentially expressed (abs(logFC)>0.25 and adjusted p-value<0.05) in “at least three patients”, and “showing the same direction of the change for the three patients”, when considering the contrast FANCA^+^ vs FANCA^-^ (n=152; see the entire list in Table S2). FANCA was excluded from the list.

To investigate if GT reprogrammed the transcriptional signature of FA HSPCs towards the one corresponding to HD HSPCs, additional gene expression analyses were conducted in HD and FA HSPCs, focusing on the 152 genes that showed significant expression changes between FANCA^+^ and FANCA^-^ HSPCs in GT treated patients (See **Figure 2E** and **Table S2**). In these studies, three different comparisons were performed in each of the twelve HSPC subpopulations: FANCA^+^ *vs*. FANCA^-^ HSPCs from GT-treated FA patients (GT-FA HSPCs); HD HSPCs *vs*. FANCA^-^ GT-FA HSPCs; and finally, HD HSPCs *vs*. FANCA^+^ GT-FA HSPCs **(Fig. 3C)**. As observed in analyses of **Fig. 2**, significant differences (dark red or blue spots in **Fig. 3C**) were most evident in HSPC subpopulations with a higher representation (i.e. monocyte CD34^+^ cells; p-value<0.001).

Regarding the comparison of GT-FANCA^+^ *vs*. GT-FANCA^-^ HSPCs (**Fig. 3C** first column), several differentially expressed genes with relevance in FA have been marked (see right side of the figure). Interestingly, genes involved in functions such as DNA repair or cell cycle were upregulated in GT-FANCA^+^ vs GT-FANCA^-^ cells. In the second column of **Fig. 3C**, the gene expression pattern of HSPCs from HDs was compared with that corresponding to uncorrected HSPCs from GT-treated patients (HD vs GT-FANCA^-^). Strikingly, most of the transcriptional changes observed between these populations resembled changes seen between corrected and uncorrected FA HSPCs (first column in **Fig. 3C**) (see **Table S3** for detailed analyses per cell type using two complementary statistical analyses). Finally, in contrast to the above-mentioned observations, the comparison of the transcriptional program of HD HSPCs vs FANCA^+^ HSPCs from GT-treated FA patients (third column in **Fig 3C**) showed limited changes, most of which were not significant and not related to differences noted when HD or corrected FA HSPCs were compared with uncorrected FA HSPCs. Overall, these studies demonstrate that lentiviral-mediated GT reverts the gene expression program of FA HSPCs, which then acquire an expression pattern that resembles the signature characteristic of healthy HSPCs.

### Lentiviral-mediated gene therapy reverts molecular pathways characteristic of FA HSPCs

Having observed that the ectopic expression of *FANCA* in FA HSPCs reverts the transcriptional signature of these cells to resemble a healthy HSPC signature, we next performed a gene-set enrichment analysis to determine changes in relevant pathways associated with FA. These include pathways related to DNA damage response and repair, cell cycle checkpoint, cell aging, and telomerase maintenance (see materials and methods). As shown in **Fig. 4** and **Table S4**, an upregulated expression of several of these pathways was observed when FANCA^+^ and FANCA^-^ HSPCs present in each of the GT-treated patients were compared (See first paired columns in **Fig. 4**; GT-FANCA^+^ vs GT-FANCA^-^). Moreover, an almost identical upregulation of these pathways was observed when HSPCs from HDs were compared with FANCA^-^ HSPCs from GT-treated patients (HD vs GT-FANCA^-^; second paired columns in **Fig. 4**).

**Figure 4.**
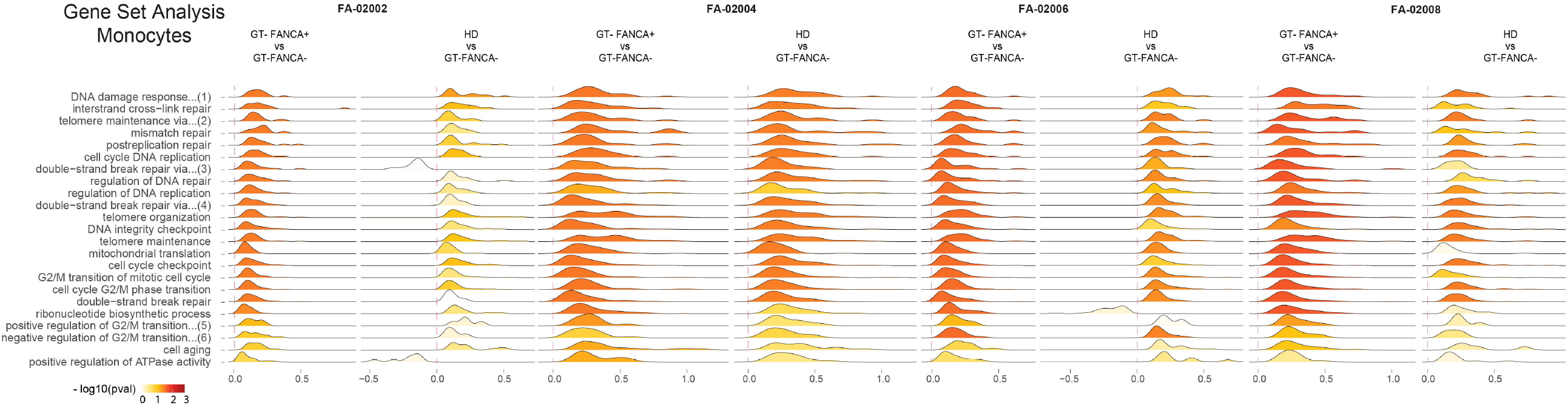
Restored gene expression pathways associated to the ectopic expression of FANCA in HSPCs from gene therapy treated FA patients. Comparative density plots showing the distribution of differential expression derived logFC for a selected set of pathways. For each patient, paired comparisons between FANCA^+^ vs FANCA^-^ HSPCs from GT-treated FA patients and HD HSPCs vs FANCA^-^ HSPCs are shown. The complete name of the pathways with numbers are: (1) DNA damage response, detection of DNA damage, (2) telomere maintenance via semi-conservative replication, (3) double-strand break repair via homologous recombination, (4) double-strand break repair via nonhomologous end joining, (5) positive regulation of G2/M transition of mitotic cell cycle, and (6) negative regulation of G2/M transition of mitotic cell cycle.

A deeper comparative expression analysis of genes involved in the cell cycle control was then performed between FANCA^+^ and FANCA^-^ HSPCs from GT-treated patients (GT-FANCA^+^ vs GT-FANCA^-^; see external crowns in **Fig. 5A**). Similar comparisons were also made between HD HSPCs and FANCA^-^ HSPCs from GT-treated patients (internal crowns in **Fig. 5A**). As shown, the ectopic expression of FANCA was associated with the downregulation of *TGF-β* in every patient, and also of *p21* (*CDKNA1*) in two of the four GT-treated FA patients. On the other hand, a number of cyclins and mini-chromosome maintenance (MCM) genes were upregulated in corrected vs uncorrected FA HSPCs (FANCA^+^ vs FANCA^-^ HSPCs). Notably, when similar comparisons were performed between HD HSPCs and FANCA^-^ HSPCs from GT-treated FA patients, almost identical gene expression changes were observed (**Fig. 5A)**, indicating that the behavior of cell cycle and DNA checkpoint pathways in corrected FA-A HSPCs resembled the pathways characteristic of healthy HSPCs.

**Figure 5.**
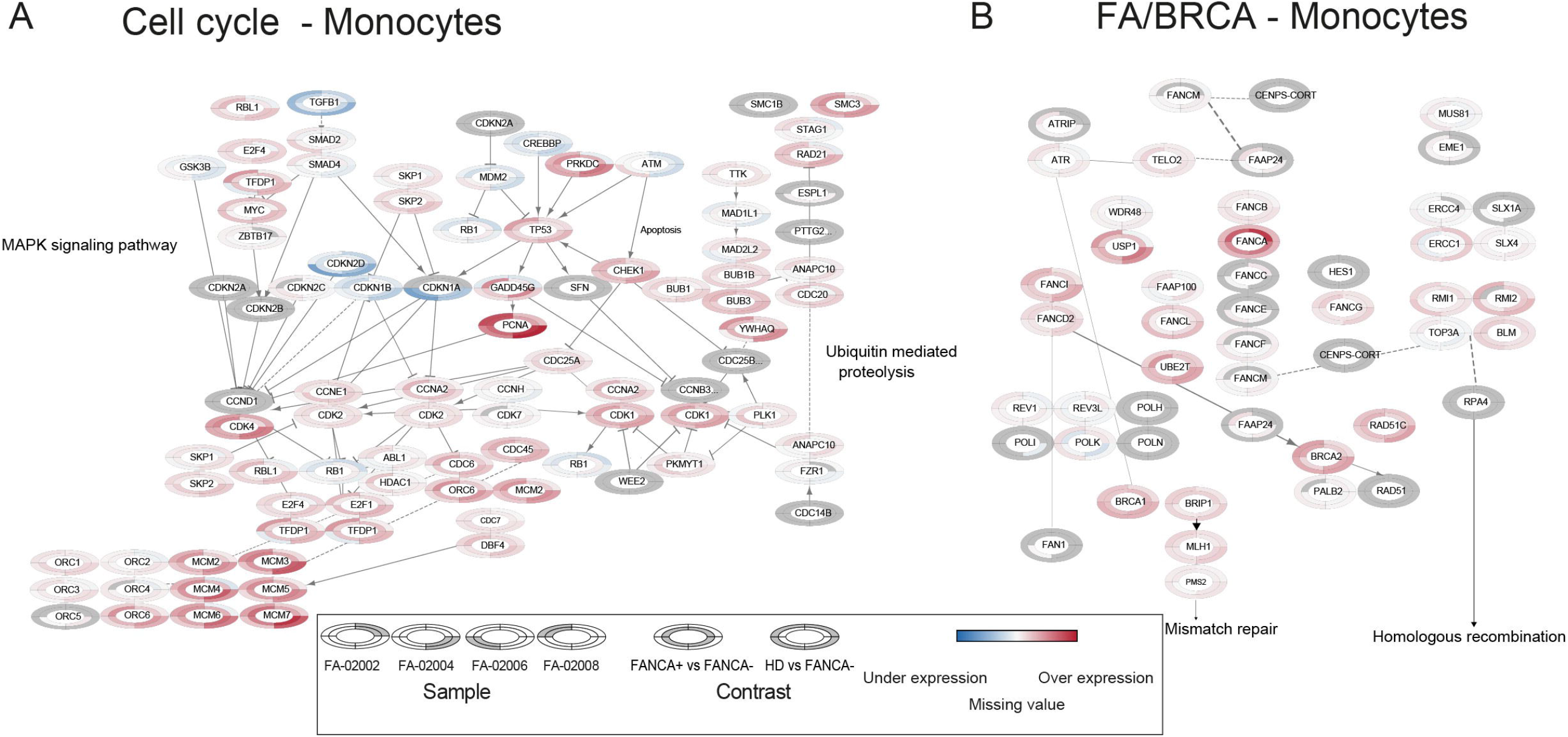
Restored gene expression of key pathways in corrected HSPCs from gene therapy treated FA patients. **(A)** Upregulated and downregulated genes implicated in cell cycle control. The figure represents the logFC associated to two contrasts: FANCA^+^ vs FANCA^-^ HSPCs from GT-treated FA patients (internal crowns), and HD HSPCs vs FANCA^-^ HSPCs from GT-treated patients (external crowns). Each external and internal crown is divided in four parts, each of them representing one FA patient. Upregulated genes are shown in red and downregulated genes in blue. **(B)** Same representation as in A showing changes in the expression of genes participating in the FA/BRCA pathway.

Since previous studies have shown that p21 participates in the transcriptional repression of different genes of the FA/BRCA pathway (Jaber, Toufektchan et al., 2016, Rego, Harney et al., 2012) changes in the expression of FA/BRCA genes were also comparatively investigated in FANCA^+^ and FANCA^-^ HSPCs from GT-treated patients. As shown in **Fig. 5B**, in addition to *FANCA*, several other genes participating in the FA/BRCA pathway, including *FANCB, FANCI, FANCD2, FANCG, UB2T* (*FANCT*), *BRCA2, PALB2, BRCA1* and *BRIP1* (*FANCJ*) were upregulated in GT-FANCA^+^ vs. GT-FANCA^-^ HSPCs. Again, many of these genes were also upregulated when HD HSPCs were compared with uncorrected GT-FANCA^-^ HSPCs.

This set of studies thus demonstrate that GT reverts physiologically relevant molecular pathways that are severely affected in HSPCs from FA patients

### Functional implications of the restored transcriptional program of corrected FA-HSPCs

In a final set of experiments, we investigated the functional implications associated with the restored transcriptional program of gene corrected FA HSPCs. First, we investigated if the restored DNA repair pathways had a direct implication in the correction of the hypersensitivity of FA-HSPCs to inter-strand crosslinking agents such as mitomycin C (MMC). While colony forming cells (CFCs) from the four patients were highly sensitive to MMC prior to the infusion of corrected HSPCs (no colonies were generated when cells were exposed to 10 nM MMC), survival of these cells to MMC was markedly increased 2 to 5 years after GT (22.3% to 67.3% of CFCs survived in 10 nM MMC; **Figure 6A**). Consistent with these studies, the percentage of T cells with diepoxybutane (DEB)-induced aberrant chromosomes was also markedly decreased after GT of FA-A patients (Figure 6B). Overall, these studies confirm the functional implications associated with the restoration of the DNA repair pathways observed in the scRNAseq analyses shown in **Fig. 4**.

**Figure 6:** Functional implications associated to the restoration of DNA repair and telomere biology pathways. **(A)** Survival to mitomycin C (MMC; 10 nM) of BM colony forming cells (CFCs) from FA-A patients shown in Figures 1-5 prior and after infusion of gene corrected cells. **(B)** Percentage of PB T cells with chromosomal aberrations induced by diepoxibutane (DEB), prior to and after gene therapy. **(C)** Analysis of the telomere length of PB cells from FA-A patients shown in Figures 1-5 at 15^th^ day post-infusion, when corrected cells were still undetectable, to the 2^nd^ −5^th^year post-infusion of gene corrected cells. As a negative control, three FA patients with no significant engraftment of gene corrected cells are included. Dashed lines correspond to telomere lengths corresponding to HD age matched PB cells. Time-points of the MMC, DEB and telomere analyses are the same or the closest ones corresponding to the scRNAseq analyses shown in Figures 1-5.

Finally, since scRNAseq studies also showed the restoration of telomere maintenance pathways, we studied changes in the telomere length of PB cells from the four FA patients included in this study. In particular, we evaluated changes that took place from the 15^th^ day post-infusion, when the proportion of corrected cells was still undetectable, to the 2^nd^ −5^th^ year post-infusion. As a negative control, changes in the telomere length of PB cells from three FA-A patients with no significant engraftment of corrected cells (VCN<0.008; control FA group) were investigated at comparable time points. While a clear reduction in the telomere length was observed in PB cells from the negative control group, a stabilization or even elongation was observed in GT treated patients 2-5 years after infusion of corrected cells. Also of interest is the comparison of the telomere length of PB cells from HDs of the same age, which indicated that telomeres from GT-treated patients were still below values corresponding to HDs.

Overall, the results obtained in this study demonstrate for the first time that GT - in particular the lentiviral-mediated GT in FA patients - reverts the transcriptional program of diseased HSPCs, which then display gene expression signatures, molecular pathways and cellular phenotypes which then resemble the physiological status of healthy HSPCs.

## DISCUSSION

Gene therapy has emerged as a safe and efficient therapeutic option for diverse monogenic disorders affecting the hematopoietic system (See review in (Ferrari et al., 2021)). In a previous study we showed for the first time preliminary clinical evidence of safety and efficacy of GT in a more complex genetic disease, such as FA (Rio et al., 2019). In that clinical study we demonstrated that the ectopic expression of FANCA reproducibly conferred engraftment and proliferative advantage of corrected HSCs in non-conditioned FA-A patients. Nonetheless, the question of whether corrected FA HSPCs acquire the transcriptional profile characteristic of healthy HSPCs was not addressed in our previous study, nor in any other GT trial of a monogenic disease.

By leveraging scRNAseq profiling we have extended the conclusions from our previous study towards the evaluation of changes in the transcriptional program of corrected versus uncorrected HSPCs that coexist in the BM of the same patient. These analyses and also gene expression comparisons performed with respect to HD HSPCs allowed us to demonstrate for the first time that GT not only corrects the expression of the mutated gene (*FANCA* in this case), but also reprograms the molecular circuits of diseased FA-A HSPCs, which then acquire a normalized gene expression pattern characteristic of healthy HSPCs.

Importantly, the restored expression of *FANCA* decreased the expression of two genes with particular implications in FA, *TGF-β* and *p21*. Increased mRNA levels of these two genes have been observed in HSPCs from FA patients and murine FA models and are believed to contribute to the BMF characteristic of the disease (Ceccaldi, Parmar et al., 2012, Zhang, Kozono et al., 2016). Notably, previous studies have shown that p21 downregulates *USP1* upon exposure to DNA damage, disrupting FANCD2/L mono-ubiquitination and nuclear foci formation (Rego et al., 2012). Additional studies have also shown that several genes involved in the telomere biology and in the FA/BRCA pathway are downregulated via the p21/E2F4 pathway (Jaber et al., 2016). Although it might be expected that p53 and Myc – which have been shown to be upregulated in FA patients (Ceccaldi et al., 2012, Rodríguez, Zhang et al., 2021) – should be also downregulated in corrected FA HSPCs, our data do not support this hypothesis. Reasons accounting for these apparent unexpected observations could be related either with the enhanced expression of cell cycle genes in corrected FA HSPCs, with post-transcriptional regulatory mechanisms involved in p53 expression (Ceccaldi et al., 2012), or also with the moderate BMF status of the FA patients enrolled in the FANCOLEN I trial (Rio et al., 2019).

Our study also shows that several defective pathways implicated in the FA cellular phenotype were significantly upregulated as a result of the ectopic expression of *FANCA* in FA-A HSPCs. Among all, the restoration of DNA repair related pathways has enormous implications in FA GT since this accounts for the correction of the hypersensitivity of BM progenitors and PB T cells to genotoxic drugs, such as MMC and DEB.

Also of note was the observation of the increased expression of mini-chromosome maintenance (MCM) helicase genes in corrected FA-A HSPCs. Previous studies have shown that decreased levels of MCM proteins are associated with the HSC hypersensitivity to replication stressors (Flach, Bakker et al., 2014), and have shown a relationship between FA and accelerated ageing (Brosh, Bellani et al., 2017). Strikingly, our study shows for the first time that corrected FA-A HSPCs upregulates the expression of MCM, emulating the MCM expression observed in HD HSPCs, and suggesting that genetic correction should reduce the replication stress and contribute to the rejuvenation of the HSCs from FA patients. Importantly, our study has also shown that GT upregulates several genes associated with the telomere biology, and revealed for the first time the maintenance or even elongation of the telomere length in PB cells from FA patients after 2-5 years post-GT. This observation contrasts with the progressive telomere attrition previously characterized in FA patients (Savage & Alter, 2008), and also with the negative FA control group included in our study. It is currently unknown whether GT will prevent the progressive telomere attrition characteristic of the disease, or even extend the telomere length of corrected hematopoietic cells to levels observed in HD cells.

Taken together, our study demonstrates for the first time that GT reverts the transcriptional program and defective molecular pathways in corrected FA HSPCs. This observation has a particular impact in FA and possibly in other HSC diseases characterized by DNA repair defects, since here we demonstrate that GT restores the molecular pathways of FA HSPCs towards a signature characteristic of healthy HSPCs.

## MATERIAL AND METHODS

### Patients and Healthy donors

FA patients included in this study, FA-02002, FA-02004, FA-02006 and FA-02008, were FA-A patients enrolled in the FANCOLEN-1 gene therapy trial (FANCOLEN-1; ClinicalTrials.gov, NCT03157804; European Clinical Trials Database, 2011-006100-12) and complied with all relevant ethical regulations approved by the ethics committees at Hospital Vall d’Hebron in Barcelona and Hospital del Niño Jesús in Madrid.

Patients were infused with autologous CD34^+^ cells after transduction with the therapeutic PGK-*FANCA*.Wpre* LV (Gonzalez-Murillo, Lozano et al., 2010). Recent clinical data has shown that although no conditioning was given to these patients prior to cell infusion, a progressive engraftment of corrected cells took place over time, implying the presence of a chimeric population of corrected and uncorrected cells in their hematopoietic tissues (Rio et al., 2019). For the participation of healthy donors (HDs) in this study (median age = 20 y/o), BM aspiration was performed after provision of informed consent, which was approved by the clinical research ethics committee of Clínica Universidad de Navarra.

### Bone marrow cells

Bone marrow cells from GT treated patients were obtained in the course of routine follow-up studies of the GT trial and as part of additional exploratory studies. Samples used in these studies were obtained 5 years (FA-02002), 4 years (FA-02004), 3 years (FA-02006) and 2 years (FA-02008) after infusion of transduced CD34^+^ cells, respectively. Patients FA-02004 and FA-02008 had been treated with eltrombopag to stimulate hematopoiesis 12 and 6 months prior to the evaluation of BM cells, respectively. In all instances samples were processed immediately after BM aspiration.

For the purification of CD34^+^ cells, erythrocytes were lysed with ammonium chloride lysis solution (0.155 mmol/L NH_4_Cl + 0.01 mmol/L KHCO_3_ + 10^−4^ mmol/L EDTA), washed using PBS (Gibco) + 0.2%BSA (10%) + 2% PenStrep (Gibco) and stained using CD45 APC (clone 2D1; Biolegend) and CD34 PECY7 (clone 4H11; eBiosciences) for 30 min at 4°C. DAPI was used at a concentration of 1 μg/mL as a viability marker. CD34^+^ cells were then sorted in a BD INFLUX™ (BD Biosciences) or BD FACSAria II (BD Biosciencs) as previously shown (Rio et al., 2019). Purified CD34^+^ cells were directly used for scRNA seq analysis and small aliquots stored at −80°C for vector copy number analysis.

### Analysis of lentiviral vector copy numbers in total BM and purified CD34^+^ cells

The number of proviral copies per cell (VCN/cell) was analyzed after genomic extraction of the DNA using the DNA easy blood and tissue kit (Qiagen) or by proteinase K lysis as previously described(Rio et al., 2019). Duplex qPCR was conducted to detect the *Psi* sequence of the provirus and the *Albumin*, as a control gene. To amplify *Psi* sequence: *Psi* forward (*Psi*.F): 5’ CAGGACTCGGCTTGCTGAAG 3’ and *Psi* reverse (Psi.R): 5’ TCCCCCGCTTAATACTGACG 3’ primers were used and detected with the Taqman probe *Psi*.P FAM: 5’CGCACGGCAAGAGGCGAGG3’. To normalize to endogenous *Albumin*, specific primers for *Albumin* were used: *Alb* forward (*Alb*.F): 5’ GCTGTCATCTCTTGTGGGCTG 3’ and *Alb* reverse (*Alb*.R.): 5’ ACTCATGGGAGCTGCTGGTTC 3’ together with a Taqman probe *Alb*.P VIC: 5’ CCTGTCATGCCCACACAAATCTCTCC 3’. qPCR was conducted in an Applied 7500 Fast Real Time PCR system (Thermo Fisher Scientific), as previously described (Rio et al., 2019).

### Single-cell RNA-sequencing (scRNA-seq)

The transcriptome of BM CD34^+^ cells was investigated using NEXTGEM Single Cell 3’ Reagent Kits v3.1 (10X Genomics) according to the manufacturer’s instructions. Between 2.000 and 6.000 CD34^+^ cells were loaded at a concentration of 700-1,000 cells/µL on a Chromium Controller instrument (10X Genomics) to generate single-cell gel bead-in-emulsions (GEMs). In this step, each cell was encapsulated with primers containing a fixed Illumina Read 1 used to sequence a cell-identifying 16 bp 10X barcode for each cell and a 12 bp Unique Molecular Identifier (UMI) for each transcript. Upon cell lysis, reverse transcription yielded full-length, barcoded cDNA. This cDNA was then released from the GEMs, PCR-amplified and purified with magnetic beads (SPRIselect, Beckman Coulter). Enzymatic Fragmentation and Size Selection was used to optimize cDNA size prior to library construction. Fragmented cDNA was then end-repaired, A-tailed and ligated to Illumina adaptors. A final PCR-amplification with barcoded primers allowed sample indexing. Library quality control and quantification was performed using Qubit 3.0 Fluorometer (Life Technologies) and Agilent’s 4200 TapeStation System (Agilent), respectively. Sequencing was performed in a NextSeq500 (Illumina) (Read1: 28 cycles; Read 55 cycles; i7 index: 8 cycles) at an average depth of 20,000 reads/cell. According to these analyses CD34^+^ cell populations were classified as *corrected* (FANCA^+^) and *uncorrected* (FANCA^-^) cells, considering that FANCA^-^ is enriched with cells that only express the endogenous mutated *FANCA* mRNA, while FANCA^+^, that includes cells with higher FANCA expression, is enriched with cells that express both the endogenous mutated *FANCA* plus the ectopic functional *FANCA* mRNA.

### scRNA-seq: bioinformatics

#### Data filtering and normalization

Sequenced libraries were demultiplexed, aligned to human transcriptome (hg38) and quantified using Cell Ranger (v_3.0.1). Ongoing analysis was conducted using Seurat (V_3.2.0)(Stuart, Butler et al., 2019) in R (V_3.5.2) (Team, 2019). Quality control filters based on the number of detected genes, number of UMIs and percentage of mitochondrial UMIs were performed to each one of the samples. The thresholds were defined based on the distribution of the previously mentioned parameters and visual inspection of quality control scatter plots. After filtering of low quality cells, a total number of 14,208 (FA-02002), 720 (FA-02004), 1,438 (FA-02006), 1,995 (FA-02008), and 12,549 (HD) cells were retained.

Each single cell dataset was individually normalized, using the Normalize Seurat function. Feature counts for each cell were divided by the total counts for that cell and multiplied by the scale factor. This was then natural-log transformed. The data was regressed out by cell cycle stadium, number of features and number of counts. Non-linear dimensional reduction was performed (UMAP) to plot the data of each sample. PCA was defined as dimensional reduction to use in the UMAP graph. Each of the FA samples was integrated with the healthy donor sample.

#### Cell annotation

For cell annotation, we use the annotation conducted in three additional human samples of healthy young individuals (3YI). The isolation protocol of 3YI includes the cell types in the 4 FA patients and the HD: Ficoll-Paque Plus (GE healthcare) density gradient centrifugation and stained using CD34 (clone 8G12; BD bioscience) CD64 (clone 10.1; Biolegend) CD19 (clone SJ25C1; Biolegend) CD10 (clone HI10A; Biolegend) CD3 (clone OKT3; Biolegend) CD36 (clone CLB-IVC7; Sanquin Plesmanlaan) CD61 (clone RUU-PL7F12; BD bioscience) for 15 min at RT. And finally, CD34+ CD64-CD19-CD10-CD3-CD36+CD61+ cells were then sorted in a BD FACSAria II (BD Biosciences) as previously shown. The data-analysis processing of those samples was conducted as the protocol described for FA samples. Next, we performed unsupervised clustering with the Louvain algorithm as implemented in Seurat (Blondel, Guillaume et al., 2008). We tested several resolution values and assessed the results by calculating the average silhouette for each cluster. We determined the cluster markers using the Seurat function FindAllMarkers, with the MAST method. Finally, we annotated the clusters in 3YI by manually inspecting the most specific markers and looking for curated markers in the literature. Using the robust annotation conducted in 3YI, the “label transfer function” from Seurat was used to annotate the four FA and the HD samples (Stuart et al., 2019). It is important to note that while the annotation of the 3YI is valid for the annotation of FA and HD samples, we decided not to include the 3YI samples in the analysis as the cell proportions may be different based on the isolation protocol.

#### Differential expression analysis

The differential expression analysis was conducted using FindMarkers function in the Seurat package. Genes were considered differentially expressed if |logFC|>0.25 and adjusted p-value<0.05.

#### GSEA analysis

Gene set enrichment analyses were conducted using ClusterProfiller (version 3.10.1)(Blondel et al., 2008) in R(Team, 2019). The normalized data of each sample and cell type was ranked by the logFC value and the analysis was run comparing our data with GO biological processes. A gene set was considered significantly enriched if GO adjusted p-value<0.05.

### Pathway visualization

After GSEA analysis two core pathways were selected (Cell cycle and FA/BRCA) for visualization purpose using Cytoscape (version 3.8.2.) (Shannon, Markiel et al., 2003). The values of logFC were for each one of the samples and different contrast using omic visualizer package. The pathways are shown as imported in the Cytoscape package; for two genes an alternative gene symbol is shown (MHF, RPA).

### Analysis of the sensitivity of hematopoietic colony forming cells to the genotoxic agent mitomycin C

To assess the influence that gene therapy had in the response of FA hematopoietic progenitors to mitomycin C (MMC), the number of colonies generated in the absence and the presence of this agent was assessed. In these experiments a total number of 2.5×10^5^ BM cells were plated in plates containing 1 mL methylcellulose medium (MethocultTM #H4434) supplemented with 10 µg/mL anti-TNFα and 1 mM N-acetylcysteine, in the absence and the presence of 10 nM MMC (Sigma-Aldrich). Cells were then cultured for 14 days at 37°C, 5% CO_2_ and 5% O_2_, and colonies were then scored under an inverted microscope.

### Chromosomal instability test in peripheral blood T cells

The diepoxybutane (DEB)-induced chromosomal instability test was carried out in peripheral blood (PB) T cells from the FA patients prior to and in the long-term after infusion of corrected cells. After 24h of culture, PB cells were incubated in the absence or the presence of DEB (0.1µg/mL; Sigma-Aldrich), and forty-six hours later colcemid was added (Gibco; 0.1 µg/mL). Two hours later, metaphase spreads were obtained and stained with Giemsa. Fifty metaphases were analyzed from DEB-treated and 25 metaphases from unexposed cultures in a Zeiss Imager M1 microscope coupled to a computer assisted metaphase finder (Metasystems). Criteria for the determination and quantification of chromosome breakage were previously described (Castella, Pujol et al., 2011).

### Telomere length studies

DNA was extracted from patient’s blood samples and the telomere length was determined by quantitative PCR as previously described (Cawthon, 2002). In this method the amount of telomere DNA (T) and of the single copy *36B4* reference gene (S) were determined by quantitative PCR for each blood sample. The ratio between these two parameters (T/S) was a measure of the relative telomere length. A control DNA isolated from the cultured cell line MCF-7 was used as an internal control in each experiment to normalize the T/S ratio obtained for the experimental samples. The telomere length of each sample was calculated from the normalized T/S ratios using the formula: telomere length in Kbp = T/S x 3.86 + 1.89. Three independent experiments with triplicates were conducted for each sample.

### Basic statistical tests

#### Proportion test

In each FA sample and cell type, a two-proportion statistical test was conducted to investigate significant differences in cell type proportion between FANCA^+^ and FANCA^-^ cells in each CD34^+^ subpopulation.

#### Annova test

We conducted a two-tail ANOVA test to investigate the differences of FANCA expression between therapy treated patients among the FANCA^+^ set for each CD34^+^ cell subpopulation.

#### Wilcoxon test

The comparison of the expression of the FANCA gene between FANCA^+^ cells in FA integrated samples and HDs was performed using a two-sided Wilcoxon test for each CD34^+^ cell subpopulation. In HD, only cells with >0 FANCA gene expression value were considered.

#### Binomial Test

We conducted a binomial test to investigate if the shared directionality of changes for two contrasts, “FANCA^+^ vs FANCA^-^” and “HD vs FANCA^+^”, was significantly over-represented. To this end, the genes sharing the same directionality for both contrasts were classified as 1, and 0 otherwise; the binomial was conducted considering a probability of 0.5, number of experiments of 382 and a one-tail p-value associated to values larger than the observed. The analysis was conducted separately for each cell type and sample.

#### Binary Correlation

We conducted a binary correlation analysis between the directionality of the same genes in the two contrasts, “FANCA^+^ vs FANCA^-^” and “HD vs FANCA^+^”. All the upregulated genes were classified as 1 and the downregulated as 0. A binary correlation test was conducted using R.

#### General considerations

To perform all the statistical tests R (Team, 2019) was used. In all the cases the multiple testing was addressed using Bonferroni; and for any analysis the null hypothesis was rejected if adjusted p-value <0.05.

## Supporting information

Expanded Figure 1

Expanded Figure 2

## Data Availability

The authors declare that the data supporting the findings of this study will be made available before the manuscript publication.
Also data will be made available upon request, and following GDPR guidelines, after peer-review publication.

## ACKNOWLEDGMENTS

The authors thank Ramón García-Escudero for careful reading of the manuscript and helpful discussions. The authors also thank A. de la Cal for coordinating the delivery of BM samples from patients with FA. The authors are also indebted to the patients with FA, their families and clinicians from the Fundación Anemia de Fanconi. This work was supported by grants from the European Commission’s Seventh Framework Program (HEALTH-F5-2012-305421) to the EUROFANCOLEN Consortium J.B. and J.Se; from Ministerio de Ciencia, Innovación y Universidades and Fondo Europeo de Desarrollo Regional (RTI2018-097125-B-I00 to P.R. and S.N.); from Gobierno de Navarra, Departamento de Desarrollo Económico y Empresarial (AGATA 0011-1411-2020-000010); from Instituto de Salud Carlos III (ISCIII) and co-financed by FEDER CIBERONC (CB16/12/00489 and CB16/12/00225); Redes de Investigación Cooperativa (TERCEL RD16/0011/0005) and from Consejería de Educación, Juventud y Deporte de la Comunidad de Madrid (AvanCell Project; B2017/BMD3692); from Fondo de Investigaciones Sanitarias, Instituto de Salud Carlos III, Spain, grant number PI20/0335 and co-funded by European Regional Development (FEDER) funds. M.A. was supported by a PhD Fellowship from Ministerio de Ciencia, Innovación y Universidades (FPU18/05488). CIBERER and CIBERONC are initiatives of the Instituto de Salud Carlos III and Fondo Europeo de Desarrollo Regional.

## AUTHORSHIP CONTRIBUTIONS

M.L., P.R., A.V-Z., N.P., S.N., D.A, B.F-V, R.M.J, M.A, A.U-A., R.P, L.S and D-G.C performed the experimental studies and analyzed the data. J.Se. and J.Z. provided critical materials. M.L., P.R., D-G.C, F.P. J.A.B wrote the manuscript. E.N. and J.D.S. reviewed the manuscript. P.R., S.N., D-G.C, F.P. J.A.B designed the study. All authors discussed the results and contributed to the preparation of the manuscript.

## THE PAPER EXPLAINED

### PROBLEM

HSC gene therapy of monogenic diseases has already demonstrated clinical efficacy in a variety of monogenic disorders, including HSC diseases such as Fanconi anemia (FA). Nevertheless, the question of whether or not these therapies can correct the transcriptional program of target cells, in particular of diseased HSCs, has never been addressed.

### RESULTS

In this study we demonstrate that gene therapy reverts the transcriptional signature of HSPCs from patients with FA. Remarkably, these HSCs changed their transcriptional program, to resemble the one corresponding to healthy donor HSPCs, and this is associated with the phenotypic correction of FA HSPCs.

### IMPACT

Our results show for the first time the potential of gene therapy to rescue defects in the HSPC transcriptional program from patients with inherited diseases, in this particular case in a disease characterized by profound defects in the HSCs.

## FOR MORE INFORMATION

https://anemiadefanconi.org/

http://www.fanconi.org/

## DATA SHARING STATEMENT

Data supporting the findings of this study are available at: https://www.ncbi.nlm.nih.gov/geo/query/acc.cgi?acc=GSE180536

## EXPANDED VIEW FIGURES LEGENDS

**Expanded view Figure S1: (A)** <u>Left panel</u>: UMAP plot showing the clustering analysis for CD34^+^ BM cells from the FA-02002 patient undergoing FANCA gene therapy. A total of 12 clusters were identified, spanning the different HSPC subpopulations. Identified clusters include an HSC cluster (hematopoietic stem cell; brown). Clusters with megakaryocytic-erythroid identity include MEP (erythroid-megakaryocyte progenitor; purple), Erythroid (erythroid progenitor; pink), and Basophils (basophil progenitor; light pink). Clusters with lympho-myeloid identity include LMPP (lymphoid-primed multipotent progenitor; light blue), Cycling-LMPP (blue), CLP (common lymphoid progenitor; orange), GMP1 and GMP2 (granulocyte-monocyte progenitor; light green and green), Monocytes (monocyte progenitor; red), DC (dendritic cell progenitor; nude), and PreB (B cells progenitor; light purple). Right panel: Distribution of FANCA positive cells (corrected cells; red) versus FANCA negative cells (uncorrected cells; blue). **(B, C)** Same as (A), including the analysis of the FA-02004 and FA-02008 respectively patient. **(D)** Cell type proportions for each individual separating in each case FANCA^+^ and FANCA^-^ cells.

**Expanded view Figure S2: (A)** UMAP plot showing the clustering analysis for CD34^+^ BM cells from the FA-02002 patient undergoing FANCA gene therapy and the healthy donor. A total of 12 clusters were identified, spanning the different HSPC subpopulations. Identified clusters include an HSC cluster (hematopoietic stem cell; brown). Clusters with megakaryocytic-erythroid identity include MEP (erythroid-megakaryocyte progenitor; purple), Erythroid (erythroid progenitor; pink), and Basophils (basophil progenitor; light pink). Clusters with lympho-myeloid identity include LMPP (lymphoid-primed multipotent progenitor; light blue), Cycling-LMPP (blue), CLP (common lymphoid progenitor; orange), GMP1 and GMP2 (granulocyte-monocyte progenitor; light green and green), Monocytes (monocyte progenitor; red), DC (dendritic cell progenitor; nude), and PreB (B cells progenitor; light purple). **(B, C)** Same as (A), including the analysis of the FA-02004 and FA-02008 respectively patient. **(D)** Distribution of cells classified as cells derived from the healthy donor (yellow), FANCA^+^ cells (red) and FANCA^-^ cells (blue) for FA-02002 patient. **(E, F)** Same as (D) including the analysis of the FA-02004 and FA-02008 respectively patient.

