## Appendix Table Legends for "Gene Therapy Restores the Transcriptional Program of Hematopoietic Stem Cells in Fanconi Anemia"

**EXPANDED VIEW TABLE LEGENDS**

**Appendix Table S1. Results of the statistical test comparing the proportion of FANCA^+^ vs FANCA^-^ HSPCs.** For each HSPC type and each patient’s sample the adjusted p-value is shown. Bonferroni was used to correct for multiple testing.

**Appendix Table S2.** Differential **FANCA^+^ signature derived from Fig.1F.** The genes included in the list are those that for at least one cell type are identified as differentially expressed (abs (logFC)>0.25 and adjusted p-value<0.05) in “at least three patients”, and “showing the same direction of the change for the three patients”, when considering the contrast FANCA^+^ vs FANCA^-^ HSPCs (n=152). FANCA was excluded from the analysis. For each gene the information is provided for each individual, for each contrast (FANCA^+^ vs FANCA^-^ and Healthy vs FANCA+) conducted for each cell type. There is a color/number code that denotes: 1/red upregulated and statistically significant; 0.5/light-red upregulated and not statistically significant; 0 no changes; -1/blue downregulated and statistically significant; 0.5/light-blue downregulated and not statistically significant

**Appendix Table S3. Comparison of the directionality of the following two contrasts: “FANCA^+^ vs. FANCA^-^“, and “Healthy vs. FANCA^-^“.** “# genes tot”, denotes the total number of genes per individual and cell-type considered for the analysis. For the two analysis the genes sharing the same directionality for both contrasts were classified as 1, and 0 otherwise (see Methods). “Binomial test”: the analysis was executed independently by cell type and sample. The adjusted p-value after Bonferroni multiple testing correction is provided. NA denotes that it was not possible to compute the p-value due to the limited number of cells. “# of genes” denotes the number of genes with the same directionality in both contrasts. Correlation: binary correlation.

**Appendix Table S4. Gene Set Enrichment Analysis.** Results of Gene Set Enrichment Analysis conducted for each FA patient, for cell type and for each of the contrasts of interest. Only significant results are provided (adjusted p-value<0.05).
