## Supplementary figures and images for "Gene Therapy Restores the Transcriptional Program of Hematopoietic Stem Cells in Fanconi Anemia"

### Expanded Figure 1

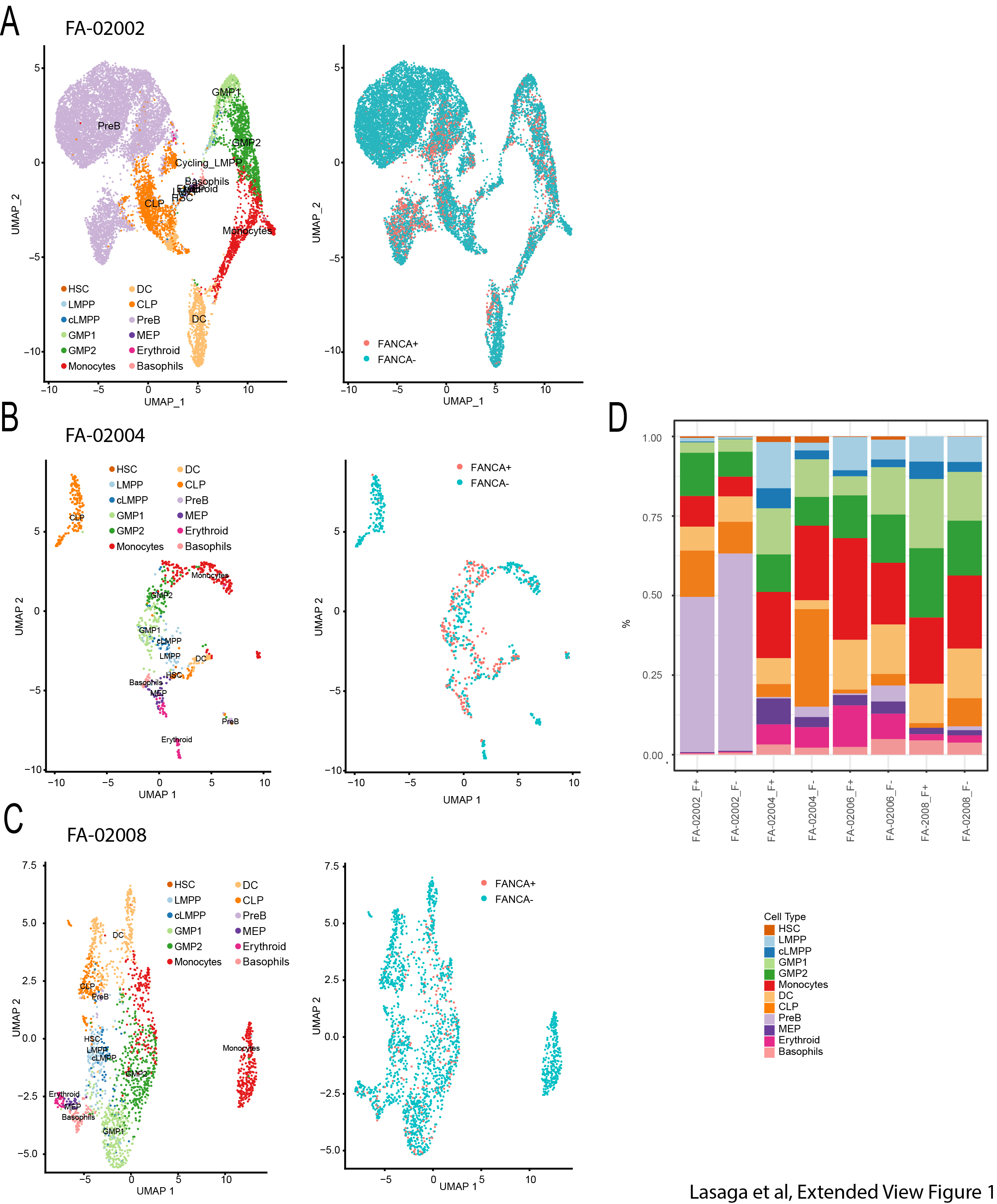

### Expanded Figure 2

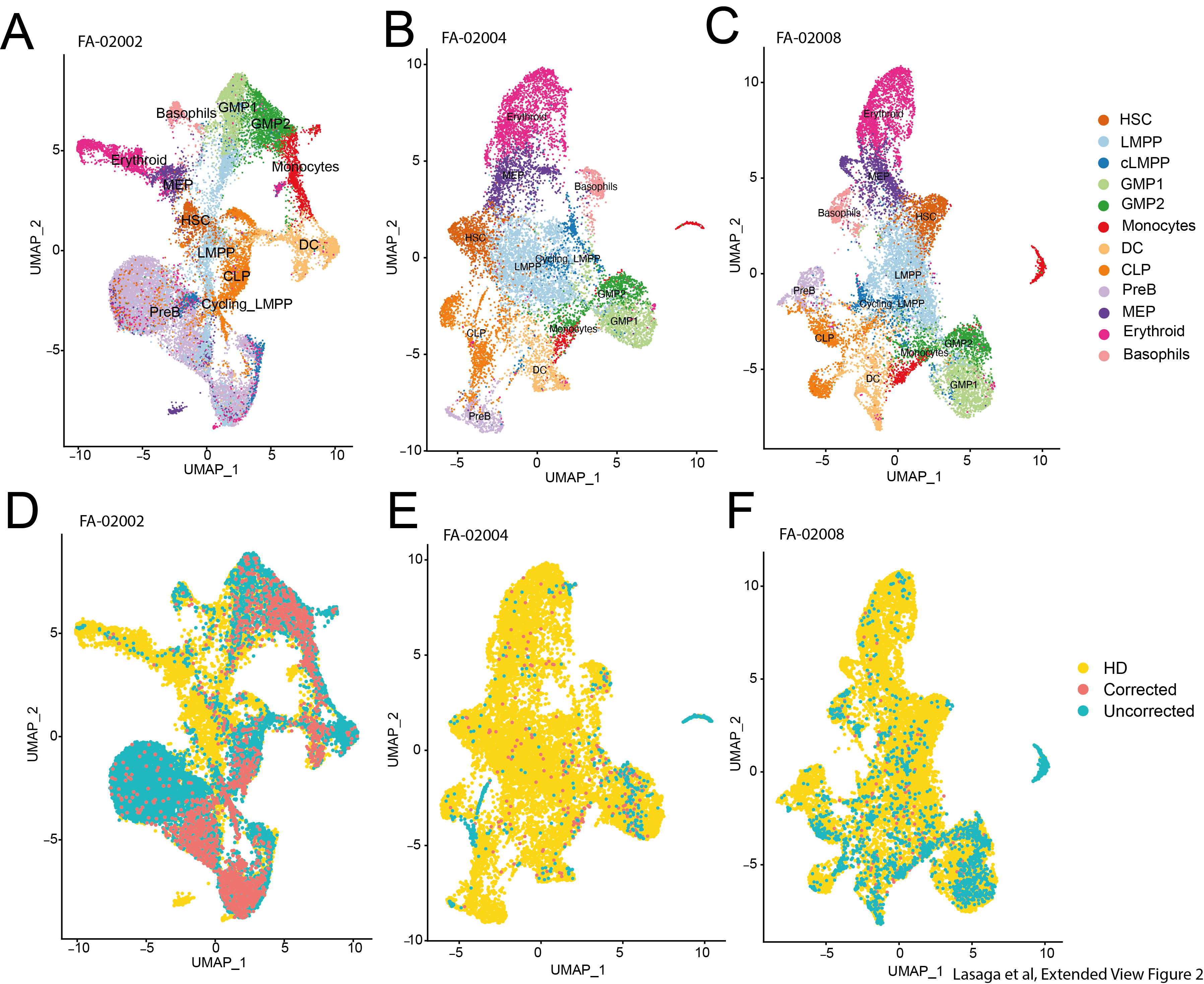
